# *Plasmodium falciparum w*ith *pfhrp2/3* deletion not detected in a 2018-2021 malaria longitudinal cohort study in Kinshasa Province, Democratic Republic of the Congo

**DOI:** 10.1101/2022.11.16.22282427

**Authors:** Ruthly François, Melchior Mwandagalirwa Kashamuka, Kristin Banek, Joseph A. Bala, Marthe Nkalani, Georges Kihuma, Joseph Atibu, Georges E. Mahilu, Kyaw L. Thwai, Ashenafi Assefa, Jeffrey A. Bailey, Rhoel R. Dinglasan, Jonathan J. Juliano, Antoinette Tshefu, Jonathan B. Parr

## Abstract

Histidine-rich protein 2- (HRP2-) based rapid diagnostic tests (RDTs) are widely used to detect *Plasmodium falciparum* in sub-Saharan Africa. Reports of parasites with *pfhrp2* and/or *pfhrp3 (pfhrp2/3)* gene deletions in Africa raise concerns about the long-term viability of HRP2-based RDTs. We evaluated changes in *pfhrp2/3* deletion prevalence over time using a 2018-2021 longitudinal study of 1,635 enrolled individuals in Kinshasa Province, Democratic Republic of the Congo (DRC). Samples collected during biannual household visits with ≥ 100 parasites/μL by quantitative real-time PCR were genotyped using a multiplex real-time PCR assay. Among 2,726 P. *falciparum* PCR-positive samples collected from 993 participants during the study period, 1,267 (46.5%) were genotyped. No *pfhrp2/3* deletions or mixed *pfhrp2/3*-intact and -deleted infections were identified in our study. *Pfhrp2/3-*deleted parasites were not detected in Kinshasa Province; ongoing use of HRP2-based RDTs is appropriate.

## Body

Progress toward malaria control and elimination in Africa requires prompt diagnosis and treatment with effective antimalarial drugs. Rapid diagnostic tests (RDTs) are widely used to identify individuals infected with *Plasmodium;* their deployment enabled significant improvements in malaria diagnostic testing across the continent over the past 15 years. The majority of RDTs used to diagnose falciparum malaria in sub-Saharan Africa detect *P. falciparum* histidine-rich protein 2 (HRP2) and its paralog HRP3, encoded by the *pfhrp2* and *pfhrp3* genes, respectively. HRP2-based RDTs are generally more sensitive and heat-stable than RDTs detecting other antigens.^1^ However, test-and-treat strategies that rely upon HRP2-based RDTs are threatened by the emergence of P. falciparum strains that escape detection due to deletion of the *pfhrp2* and/or *pfhrp3 (pfhrp2/3)* genes.^2,3^ High prevalence of *pfhrp2/3*- deleted parasites in Eritrea,^4^ Ethiopia,^5,6^ Djibouti,^7,8^ and surrounding countries recently prompted changes in malaria diagnostic testing policies. Reports from sub-Saharan countries outside of the Horn of Africa, however, indicate lower prevalence.^9^

The Democratic Republic of the Congo (DRC) has one of the highest malaria burdens in the world, accounting for 12% of global malaria cases and deaths.^2^ In the DRC, we previously reported 6.4% *pfhrp2* deletion prevalence in samples from a 2013-2014 nationally representative cross-sectional survey of asymptomatic children under five years of age.^10^ However, no *pfhrp2/3* deletions were observed in our 2017 cross-sectional study of symptomatic children and adults across three DRC provinces.^11^ These studies were both cross-sectional and did not provide information about how the prevalence of deletions may be changing over time. This study aims to estimate the *pfhrp2/3* deletion prevalence and changes over time in Kinshasa Province, DRC.

This study includes samples collected as part of a 2018-2021 longitudinal study of malaria conducted at seven sites across Kinshasa Province, DRC. A total of 1,635 participants were enrolled in one urban neighborhood, three peri-urban villages, and three rural villages **(Figure 1)**. Study visits were conducted as part of twice-yearly household surveys in the dry and rainy seasons (active surveillance) and as part of routine care at local health centers (passive surveillance) as previously described.^12,13^ At each visit, a comprehensive questionnaire on malaria symptoms and treatment, and bed net usage was administered. A finger- or heel-prick sample was obtained at each visit for RDT (SD Bioline Ag P.f./Pan RDT [05FK60], Alere, Gyeonggi-do, Republic of Korea, or CareStart, Access Bio, Somerset, U.S.A.) and dried blood spot (DBS) preparation for future molecular investigation. RDT-positive patients were treated according to national guidelines.

**Figure 1.**
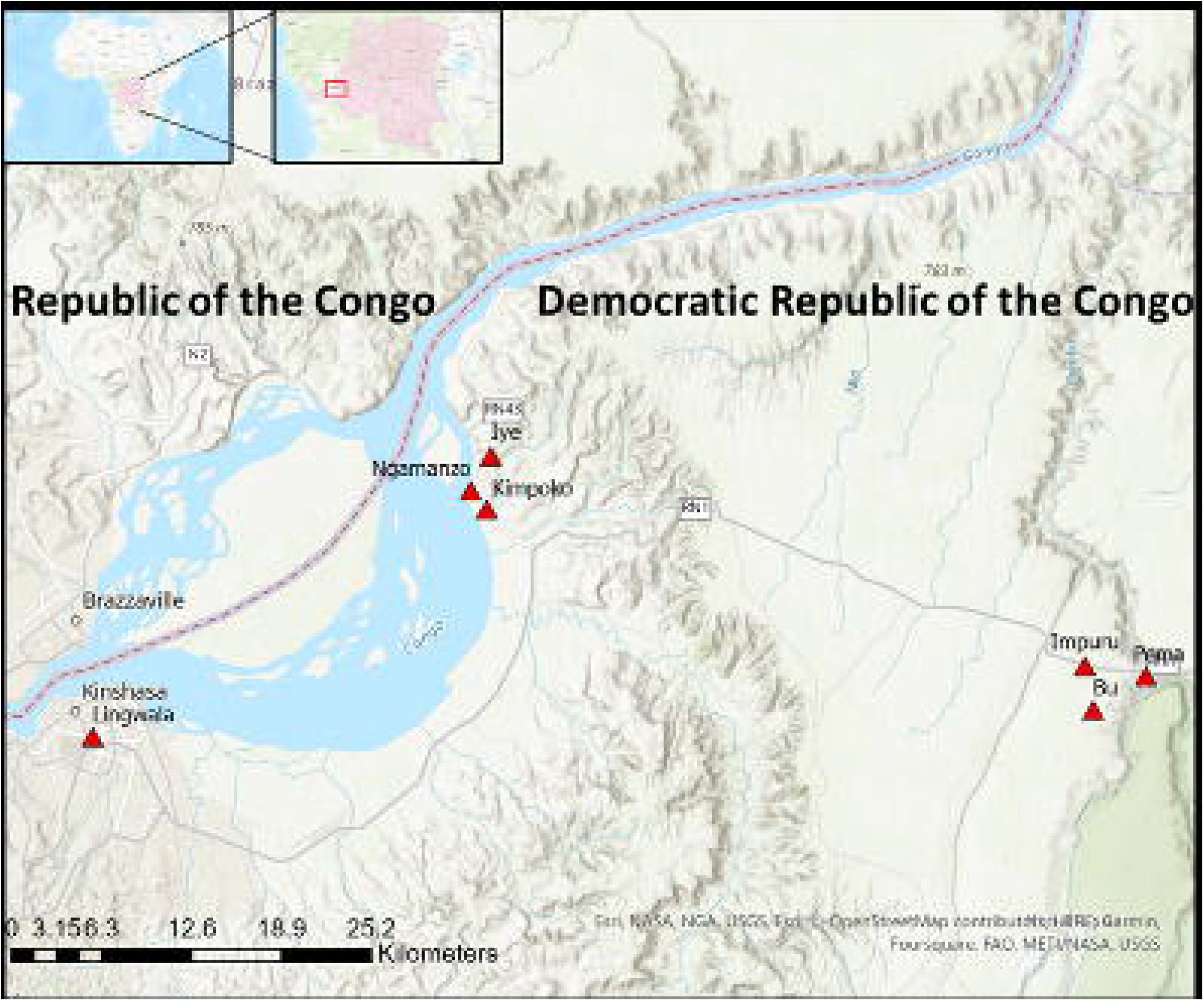
Study sites in Kinshasa Province, DRC

DNA was extracted from DBS using Chelex-100 and saponin or tween as previously described. ^14,15^ Quantitative real-time PCR (qPCR) targeting the *P. falciparum* lactate dehydrogenase (*pfldh*) gene was used to estimate P. falciparum parasitemia, using serial dilutions of DNA extracted from a mock DBS made with cultured *P. falciparum* 3D7 or FCR3 strain parasites at known parasite density. Samples with ≥ 100 parasites/μL were selected for *pfhrp2/3* deletion identification using a multiplex real-time PCR assay that detects *pfldh, pfhrp2, pfhrp3*, and *human beta-tubulin (HumTuBB)*.^16^ We used this parasite density threshold to reduce the risk of unintentional misclassification of deletions in the setting of low DNA concentrations.^17,18^ Positive calls required cycle threshold (C_t_) values < 35. Samples positive for *HumTuBB* and *pfldh* but negative for pfhrp2 or pfhrp3 were subjected to a confirmatory real-time PCR targeting the P. falciparum beta-tubulin (*PfBtubulin*) gene. Deletion calls were limited to samples positive for *HumTuBB* and both single-copy P. falciparum genes (*pfldh* and *pftubulin*), but negative for pfhrp2 and/or pfhrp3. Mixed infections of pfhrp2/3-intact and -deleted strains were defined conservatively as samples in which (*pfhrp2* C_t_ - *pfldh* C_t_) >3 or (*pfhrp3* C_t_ - *pfldh* C_t_) > 3. All assays included *P. falciparum* 3D7, DD2, and HB3 strain DNA as wildtype, *pfhrp2*-deleted, and *pfhrp3*-deleted controls, respectively. All participants provided informed consent. Ethical approval for this study was granted by the Institutional Review Boards of the University of North Carolina-Chapel Hill and the Kinshasa School of Public Health.

A total of 1,267 samples collected from 649 individuals in 179 households between 2018 and 2021 were included in this study **(Figure 2)**. Among these, the median number of *P. falciparum* infections with ≥ 100 parasites/μL detected per participant was 2.0 (IQR: 1-3). The median age at enrollment in 2018 was 9 years old (IQR: 5-15 years old); 48.8% reported female gender. At the time of enrollment, 48.7% reported malaria in the preceding 6 months, and 42.7% of those reported more than one episode in that 6-month period. The median household size was 8 (IQR: 6-10), with high bed net coverage across the study population (90.2%). The baseline characteristics of the study population are summarized in **Table 1**.

**Table 1.** Baseline characteristics of the study population included in this analysis (1,267 samples with ≥ 100 *Plasmodium falciparum* parasites/μL from 649 participants), during enrollment in the Kinshasa longitudinal study.

|  | N = 649 participants |
| --- | --- |
| Age in years, median (IQR) | 9 (5-15) |
| Age strata, n (%) |  |
| < 1 year | 15 (2.3) |
| 1-5 years | 166 (25.6) |
| 6-10 years | 191 (29.5) |
| 11-15 years | 119 (18.4) |
| 16-25 years | 76 (11.7) |
| > 25 years | 81 (12.5) |
| Sex, n (%) |  |
| Female | 317 (48.8) |
| Male | 332 (51.2) |
| Malaria in previous 6 months, n (%) |  |
| No | 311 (51.3) |
| Yes-once | 169 (27.9) |
| Yes-many | 126 (20.8) |
| Slept under bed net previous night, n (%) |  |
| No | 47 (9.8) |
| Yes | 432 (90.2) |
| Household size, median (IQR) | 8 (6-10) |
| Site of residence, n (%) |  |
| Bu (rural) | 87 (13.4) |
| Impuru (rural) | 123 (19.0) |
| Pema (rural) | 121 (18.6) |
| Ngamanzo (peri-urban) | 108 (16.6) |
| Iye (peri-urban) | 60 (9.2) |
| Kimpoko (peri-urban) | 122 (18.8) |
| Lingwala (urban) | 28 (4.3) |
| Number of samples per participant, median (IQR) | 2 (1-3) |

**Figure 2.**
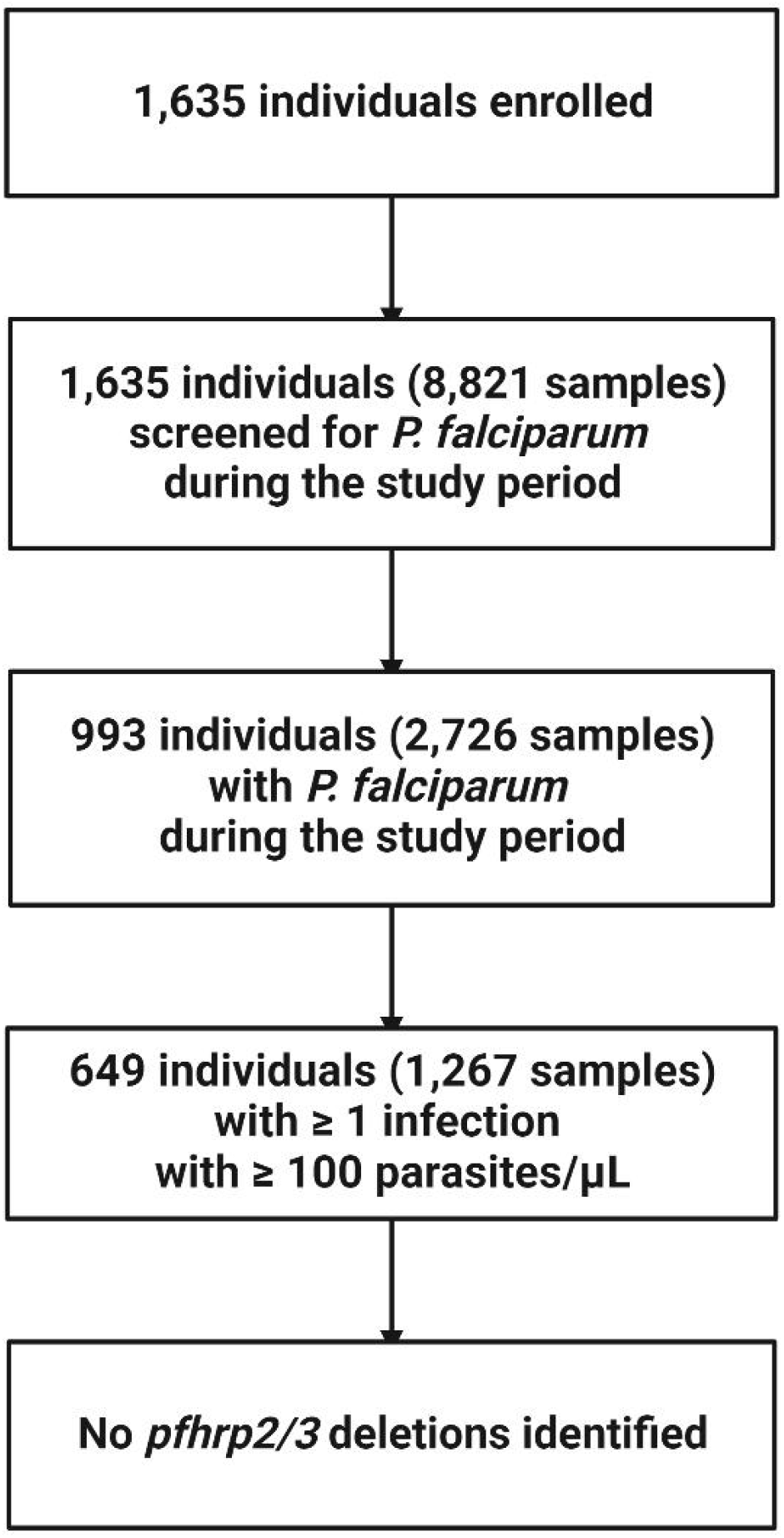
Flow diagram for the detection of *pfhrp2/3* deletion in 1,267 samples from 649 participants in phase two of the longitudinal study (2018-2021) in Kinshasa Province, DRC

All 1,267 samples had detectable human DNA as indicated by amplification of *HumTuBB* with C_t_ < 35. The multiplex PCR confirmed *P. falciparum* parasitemia in all but 2 samples which failed to amplify *pfldh*. Both *pfhrp2* and *pfhrp3* were negative in one sample (C_t_ > 35 for both gene targets). However, *pfldh* was negative in the multiplex assay and *Pftubulin* negative in follow-up testing, indicating that the original *pfldh* qPCR result was a false-positive. Thus, this sample did not meet criteria to be considered a *pfhrp2/3*-deleted parasite. There were no mixed infections of pfhrp2/3-intact and -deleted strains identified in our study population.

Overall, we did not find evidence of *pfhrp2/3* deletion in parasites sampled during our longitudinal cohort in Kinshasa Province. Our conservative approach to calling *pfhrp2/3* deletions prevents us from detecting low-density infections and thus could underestimate the true prevalence of these strains. These study results differ from our previous DRC *pfhrp2/3* deletion prevalence estimates; this discrepancy could be attributed to spatial heterogeneity in prevalence, incorrect deletion calls due to laboratory artifact, or different approaches used to identify *pfhrp2/3* deletions. However, our finding of no deletions is in line with our more recent study of symptomatic individuals and a similar study of asymptomatic and symptomatic school-aged children that showed no to little evidence of *pfhrp2/3* deletion in the DRC.^11,19^ Together, these results support the continued use of HRP2-based RDTs for the diagnosis of malaria in Kinshasa Province, DRC.

## Data Availability

All data produced in the present study are available upon reasonable request to the authors.

## Acknowledgements

The authors thank the study participants and field teams who conducted study visits. They also wish to express their gratitude to the late Prof. Steven Meshnick for mentorship and his role in the longitudinal study upon which this analysis was based. The following reagents were obtained through BEI Resources, NIAID, NIH: Genomic DNA from *P. falciparum* strain 3D7, MRA-102G, contributed by Daniel J. Carucci; *P. falciparum* strain HB3, MRA-155G, contributed by Thomas E. Wellems; *P. falciparum* strain Dd2, MRA-150G, contributed by David Walliker.

## Co-author contact information

Ruthly François: University of North Carolina at Chapel Hill, Chapel Hill, NC, United States;

Melchior Mwandagalirwa Kashamuka^2^: Kinshasa School of Public Health, Kinshasa, Democratic Republic of the Congo;

Kristin Banek^1^: University of North Carolina at Chapel Hill, Chapel Hill, NC, United States;

Joseph A. Bala^2^: Kinshasa School of Public Health, Kinshasa, Democratic Republic of the Congo;

Marthe Nkalani^2^: Kinshasa School of Public Health, Kinshasa, Democratic Republic of the Congo;

Georges Kihuma^2^: Kinshasa School of Public Health, Kinshasa, Democratic Republic of the Congo;

Joseph Atibu^2^: Kinshasa School of Public Health, Kinshasa, Democratic Republic of the Congo;

Georges E. Mahilu^2^: Kinshasa School of Public Health, Kinshasa, Democratic Republic of the Congo;

Kyaw L. Thwai^1^: University of North Carolina at Chapel Hill, Chapel Hill, NC, United States;

Ashenafi Assefa^1^: University of North Carolina at Chapel Hill, Chapel Hill, NC, United States;

Jeffrey A. Bailey^3^: Brown University, Providence, RI, United States;

Rhoel R. Dinglasan^4^: University of Florida, Gainesville, FL, United States;

Jonathan J. Juliano: University of North Carolina at Chapel Hill, Chapel Hill, NC, United States;

Antoinette Tshefu^2*^: Kinshasa School of Public Health, Kinshasa, Democratic Republic of the Congo;

Jonathan B. Parr: University of North Carolina at Chapel Hill, Chapel Hill, NC, United States;

